# Patient-Level Clinical Expertise Enhances Prostate Cancer Recurrence Predictions with Machine Learning

**DOI:** 10.1101/2022.03.22.22272635

**Authors:** Jacqueline Jil Vallon, Neil Panjwani, Xi Ling, Sushmita Vij, Sandy Srinivas, John Leppert, Mohsen Bayati, Mark K. Buyyounouski

## Abstract

With rising access to electronic health record data, application of artificial intelligence to create clinical risk prediction models has grown. A key component in designing these models is feature generation. Methods used to generate features differ in the degree of clinical expertise they deploy (from minimal to population-level to patient-level), and subsequently the extent to which they can extract reliable signals and be automated. In this work, we develop a new process that defines how to systematically implement *patient-level* clinician feature generation (CFG), which leverages clinical expertise to define concepts relevant to the outcome variable, identify each concept’s associated features, and finally extract most features on a per-patient level by manual chart review. We subsequently apply this method to identifying and extracting patient-level features predictive of cancer recurrence from progress notes for a cohort of prostate cancer patients. We evaluate the performance of the CFG process against an automated feature generation (AFG) process via natural language processing techniques. The machine learning outcome prediction model leveraging the CFG process has a mean AUC-ROC of 0.80, in comparison to the AFG model that has a mean AUC-ROC of 0.74. This relationship remains qualitatively unchanged throughout extensive sensitivity analyses. Our analyses illustrate the value of in-depth specialist reasoning in generating features from progress notes and provide a proof of concept that there is a need for new research on efficient integration of in-depth clinical expertise into feature generation for clinical risk prediction.

## 1. Introduction

Over the past decade, access to electronic health record data has become ubiquitous. As a consequence, the use of artificial intelligence (AI) to guide patient-level decision making in clinical practice has grown. One such application of AI is to create clinical risk prediction models^1–7^. These models leverage patient data to predict clinically relevant outcomes and subsequently guide allocation of medical treatments. A key component in designing these models is generating the features (i.e., covariates) supplied to the model from structured data (e.g., laboratory values) and unstructured data (e.g., physician progress notes)^8,9^. Methods used to generate features from unstructured data differ in the degree of clinical expertise they leverage, which may be as little as using specialized medical language to as much as incorporating physician-level clinical judgment^10–15^. We hypothesize the degree of clinical expertise used (minimal, population-level, and patient-level) to extract features can have a substantial impact on both the quality of the risk prediction models and the extent to which automation is possible.

In recent years, methods that generate features with *minimal* clinical expertise have gained traction. These methods rely on clinical input to identify relevant features from structured data sources, but leverage automated natural language processing (NLP) methods, some of which are pretrained on a large-scale corpus of medical text, to generate features from unstructured patient-level progress notes^16–24^. The driving force behind the rapid adoption of these approaches for feature generation is rooted in their ability to be an almost fully automated process. Advanced AI algorithms are able to tease out features from progress notes to predict an outcome described within, as well as construct new features that are often mathematically complex functions of the input features. Recent research on this automated feature generation (AFG) method, however, highlights a potential shortcoming of this method, in which it creates new biases or amplifies existing ones^4,21,25–28^.

A more classical approach to feature generation involves *population-level* clinical expertise combined with AI. This method leverages clinical feedback to create keywords and rules that can programmatically extract available features at scale for the entire patient population. The extent to which clinical expertise is utilized in this method is therefore restricted to formulating domain-specific language. Leveraging clinical knowledge from practitioners or medical databases or ontologies to create features are examples of this^29–34^. Unlike the AFG method which acts as a black-box and outputs features that have little meaning to a clinician, this method extracts features that are clinically recognizable as it relies on specially crafted medical vocabulary. However, this method requires being able to programmatically extract the features at scale. Therefore, it may overlook features that are key to the prediction task but cannot be automatically retrieved at large with a set of rules. There are numerous examples in literature in which this approach with population-level clinical expertise is actually outperformed by AFG approaches^16,35–37^.

Lastly, in this work, we sought to create a method that uses *patient-level* clinical expertise, which requires extensive clinical knowledge throughout the entire feature generation process (including model training, testing, and possibly, prospective applications), leveraging both relevant domain-specific medical language and clinical acumen. In contrast to a method with population-level clinical expertise, a patient-level clinician feature generation (CFG) approach requires substantial per-patient manual work that is not scalable to generate features from especially unstructured data sources. In particular, it leverages clinical expertise to define concepts relevant to the outcome variable, identify each concept’s associated features, and finally extract most features on a per-patient level by manual chart review. To the best of our knowledge, given the extensive patient-level reasoning required to perform such a method, a process for it has not been systematically outlined and completed, and its value measured by prediction accuracy in a clinical risk prediction model remains unclear.

In this article, we develop a systematic framework to deploy patient-level CFG and apply it to physician progress notes from Stanford Healthcare to predict the actionable outcome of 5-year prostate cancer recurrence. To assess the value of the patient-level CFG method, we implement an AFG method that extracts features from progress notes using NLP models. We choose to solely compare the patient-level CFG method to an AFG method in this paper, because the patient-level CFG method partially incorporates techniques from population-level clinical expertise methods, and as discussed above, there has been a rise in literature illustrating AFG approaches outperform population-level clinical expertise methods.

## 2. Methods

### 2.1 Collaborative Process

To begin this project, our team of clinicians and data scientists had extensive biweekly meetings for over a year, discussing the disease’s natural history, treatment options, potential patient-specific features, and clinically relevant outcome measures. This process in part entailed reading physician progress notes as a team and understanding how a clinician utilizes these notes in practice, including what information is relevant and how it is typically summarized to make day-to-day clinical decisions.

Through this exploratory work and exchange of knowledge, we discovered that, while physician progress notes include tremendous information that is leveraged in daily clinical practice to understand a patient’s disease trajectory, some features could not be extracted programmatically with a set of population-level rules (e.g., percentage core-length involvement with cancer per systematic core) and instead require extraction at a per-patient level with human reasoning. This realization motivates the project presented in this paper, as we then sought to create a systematic process to apply per-patient clinical expertise in defining and extracting patient features from progress notes.

In particular, in this project, we establish a framework for this process that at a high-level consists of two main steps: first, translating extensive knowledge learned through clinical practice and continued medical education into important domain-specific concepts required to understand the outcome of interest; and second, converting the concepts into patient-level features that can be manually retrieved from unstructured data sources with further assistance of clinical expertise. We then subsequently complete the necessary time-intensive process using data from patients who were treated for prostate cancer at the Stanford Cancer Institute between 2005-2015, under the review and approval of the institutional review board (IRB) of Stanford University.

For each patient, we extract data from the Electronic Health Records (EHR). Available patient data includes structured fields (e.g., laboratory values) and unstructured physician progress notes (e.g., biopsy reports and clinical notes). We apply minimal clinical expertise to create a set of *baseline features* from the structured data sources. For the unstructured notes, we identify and extract features through our newly developed patient-level CFG protocol. To contrast the performance of this CFG method to an AFG method, we apply automated natural language processing (NLP) methods to the progress notes for feature extraction, and subsequently compare the performance of machine learning models trained and tested on varying feature sets to establish our conclusions.

In this work, we focus on the application of the patient-level CFG process applied to progress notes. However, this process is not restricted to notes; it can be applied to other unstructured data, such as images, or even structured fields, although it is most amenable to unstructured data. While this process is generalizable, it is time-intensive, restricting this particular work to be a case study of patient-level CFG applied to only physician progress notes.

### 2.2 Patient Cohort

Inclusion criteria for the patient cohort (N=147) are: individuals who have a prostate-specific antigen (PSA) lab value available > 5 years after treatment, have a pre-treatment biopsy report with the 12 systematic cores recorded ([right/left] [apex/mid/base] [lateral/medial]), and received a prostatectomy or radiation therapy as their first treatment and had no subsequent treatment within five years. The first criterion ensures we do not misclassify patients as not having cancer recurrence due to loss to follow-up, as the outcome of cancer recurrence is computed from PSA lab values. The second criterion constrains the study to patients who have sufficient information in a pre-treatment biopsy report to capture a representative description of their disease state. We perform a sensitivity analysis that captures the timing of this biopsy report that contains the 12 systematic cores and show results are qualitatively unchanged. Finally, the third criterion is necessary because we are interested in whether a patient’s cancer recurs after their first treatment (prostatectomy or radiation therapy). This last criterion excludes an additional 21 patients who received a treatment that entailed a prostatectomy and then radiation therapy within a timespan of five years. This multimodal treatment for a patient is an alternative treatment plan, compared to only a prostatectomy or only radiation therapy, and is typically decided before any first treatment is begun. Because of this, the decision to undergo this form of treatment provides no signal as to whether the patient would have experienced cancer recurrence before the second part of their assigned treatment. Therefore, we cannot extrapolate the counterfactual had the patient only undergone their first treatment, posing difficulties in computing an unbiased estimate of the outcome for this cohort. While the above inclusion criteria may create inadvertent biases in the data, we expect such biases (if present) to not impact our comparison of different feature generation approaches and the main conclusions of the paper. Table 1 shows summary statistics for our final patient cohort.

**Table 1:** Summary statistics of patient cohort.

| N = 147 | Metric for Continuous Features |  |
| --- | --- | --- |
|  | Mean (SD) | Median |
| Age at Treatment | 68.5 (8.4) | 68.7 |
| Charlson Comorbidity Index | 5.2 (2.0) | 5.0 |
| Maximum Pre-treatment PSA | 10.8 (21.5) | 6.2 |
| Percent Positive Regions | 40.8 (26.5) | 33.3 |
| Patients with biological failure (%) | 11 (7.5) |  |
| Race = White (%) | 110 (74.8) |  |
| Staging (%) |  |  |
| t1 | 75 (51.0) |  |
| t2 | 47 (32.0) |  |
| t3 | 12 (8.2) |  |
| Grade Group of Maximum Gleason Score (%) |  |  |
| 1 | 37 (25.2) |  |
| 2 | 36 (24.5) |  |
| 3 | 31 (21.1) |  |
| 4 | 17 (11.6) |  |
| 5 | 26 (17.7) |  |

### 2.3 Outcome

All models in our analyses predict the probability of biological failure (BF) within five years after the end of treatment signaling cancer recurrence. Biochemical failure is the gold-standard for defining recurrent prostate cancer and an important endpoint because it prompts an evaluation for local recurrence and metastatic disease, and in some cases additional therapy. For patients who receive radiation therapy, a patient is classified as BF if they have a PSA level post-treatment that is at least 2 ng/mL greater than their post-treatment nadir^38^. For patients who undergo a prostatectomy, they are classified as BF if they have a PSA level of 0.4 ng/mL or above with the subsequent PSA level increasing after the end of treatment^39^. Given there is variability in the latter definition, we complete a sensitivity analysis on the results in which we classify a patient who got a prostatectomy as BF if they have a PSA level post-prostatectomy of 0.2 ng/mL or above with the subsequent PSA level also being 0.2 ng/mL or above ^40^.

### 2.4 Baseline Features

Baseline features include features from structured data sources that require minimal data analysis to compute with the output from the EHR. These include demographic and socioeconomic features, the maximum pre-treatment PSA level, and the Charlson Comorbidity Index^41^. We include the features of race, ethnicity, and language in our study, but perform a sensitivity analysis on the results excluding these three features to ensure that the outcome prediction machine learning models are not using these features in any biased fashion. For the purpose of our study, the baseline features require minimal clinical expertise to identify, either because they are readily available in the EHR (e.g., PSA values) or because there has been extensive research already conducted on how to compute them providing us with a template to follow (e.g., Charlson Comorbidity Index).

### 2.5 Patient-Level Clinician Feature Generation (CFG)

To generate the patient-level clinician features, over the span of a year from the end of 2020 to 2021, our team of oncologists (M.K.B., N.P., S.S.) and data scientists (J.J.V., M.B., S.V., X.L.) at the Stanford Cancer Institute collaborated to curate clinically relevant features from the medical record. The overarching process, as illustrated in Figure 1, leverages clinical expertise to evaluate a patient’s likelihood of prostate cancer recurrence to define the clinical concepts and their associated features that comprise the curated dataset. For example, one concept is that tumor volume in the biopsy specimen is prognostic and features representative of tumor volume include the percentage of positive regions (i.e., first grouping cores by the region of the prostate from which they were taken and then determining whether a region had any positive cores) and the percentage core-length involvement with cancer. Supplement A outlines the list of clinical concepts. All progress notes examined by the team in this step were pre-treatment, ensuring that information of the outcome variable was unknown during this process. Our team of oncologists also identified clinically relevant summary statistics that represent information both used in practice and taught to be of importance in clinical training and must therefore be generated as additional features. The result of this step is an acquired list of features that are both of clinical importance in predicting prostate cancer recurrence and available in the data.

**Figure 1:**
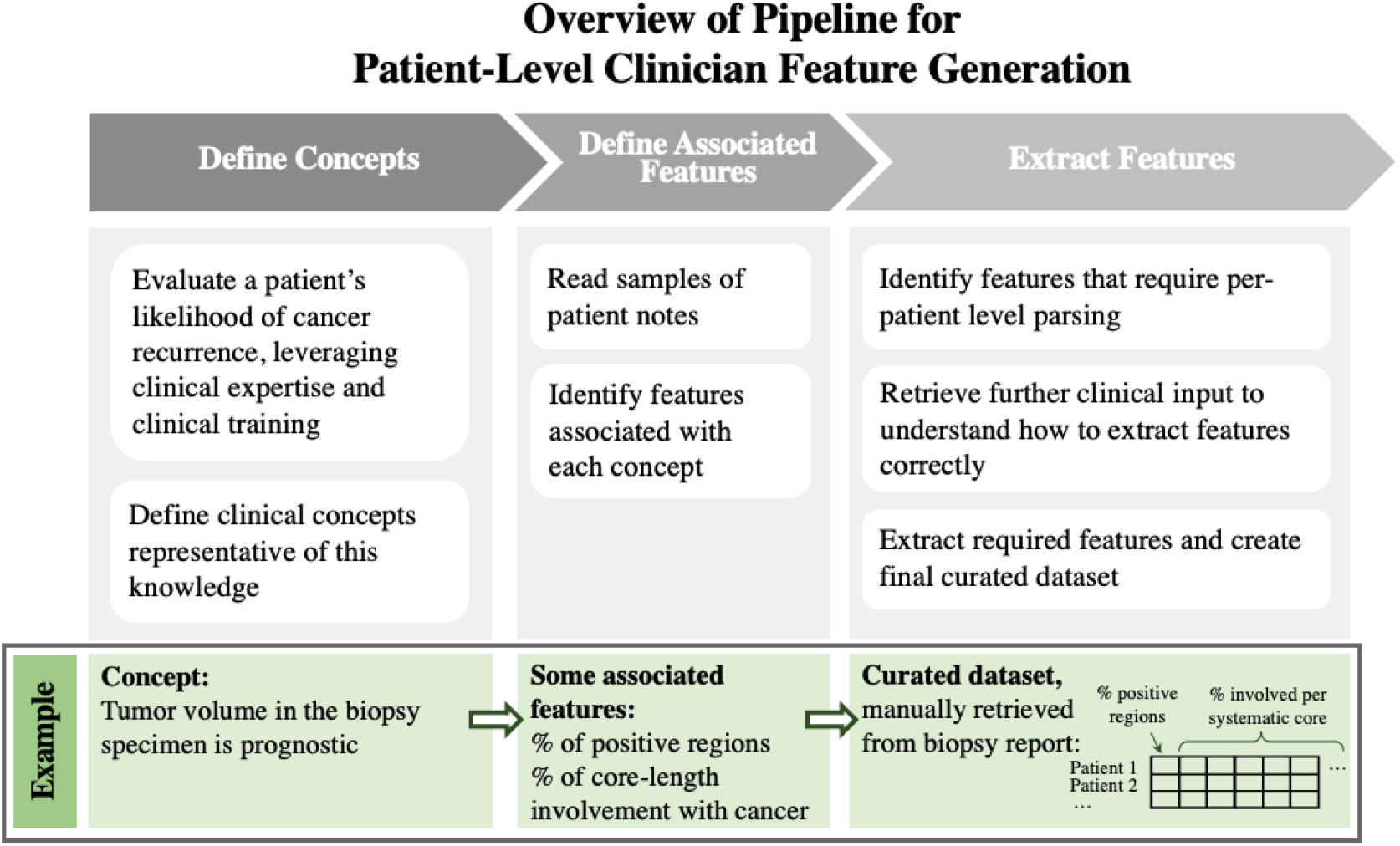
Overview of pipeline used to generate CFG features.

Then, to extract these named clinician features from notes, while a handful of them, such as the clinical T stage^42^ and maximum Gleason score, can be extracted on a cohort level leveraging regular expressions, the majority of features from the biopsy reports require manual parsing on a per-patient level because of the unstructured and unstandardized nature of the reports. This retrieval step requires even further clinical input, this time from pathologists who write the biopsy reports, to correctly understand the variable wording in the texts and to extract the features correctly. Traits of our data that contribute to the required patient-level generation of the clinician features from notes include: 1) different naming conventions for regions and different number of regions sampled; 2) distinct characters (or none at all) to separate information for each region; 3) diverse reporting of results for a region (some reports combine cores per region if they have the same results, while others keep each core strictly separate); 4) variable structures, especially for reports summarizing slides submitted from an outside institution; 5) inconsistent notation for the percent of the core involved (for example, some texts only state the length of the core involved in the diagnosis section, which must subsequently be divided by the length of the core found in the denser part of the text to compute the percent of the core involved, and this is made more difficult to do systematically with reports varying in the units used, including cm and mm); and 6) irregular wording for negated phrases such as “no adenocarcinoma” and “no significant abnormalities”. Figure 2 shows examples of each of these obstacles in real patient texts to illustrate that it is very hard to create a set of rules that could be programmatically implemented to extract these features at large.

**Figure 2:**
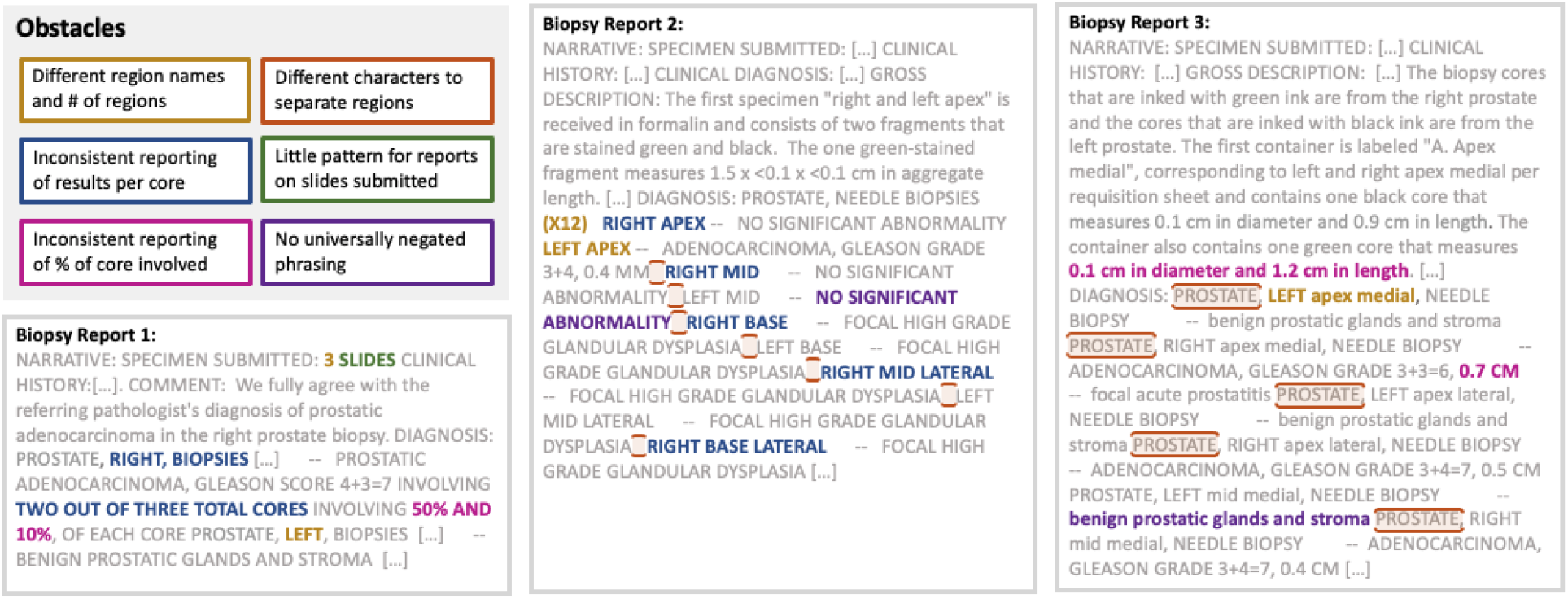
Examples of obstacles in automating the extraction of CFG features.

We denote this entire set of features using extensive patient-level clinical expertise as “CFG features”.

### 2.6 Automated Feature Generation (AFG)

To retrieve features from progress notes using an AFG method, we apply natural language processing (NLP) methods. For each patient, we create a concatenated progress note text, by combining the pre-treatment clinical note that has clinical T stage information present closest to the start date of treatment, identified using regular expressions, with the pre-treatment biopsy report from which we retrieve the subset of CFG features per systematic core (i.e., grade group, percent involved, and percent Gleason pattern 4/5 per systematic core). We then apply six different NLP methods to this concatenated text to generate features: Bag-of-Words (BoW Classic), Bag-of-Words Term Frequency-Inverse Document Frequency (BoW TF-IDF), Bidirectional Encoder Representations from Transformers (BERT), DistilBERT, ClinBERT, and Longformer.

BoW is a simple, interpretable NLP method. For the two BoW methods, we first preprocess the data to reduce the dimensionality, converting all text to lowercase, removing common English stopwords (using the package “nltk” in Python), and lemmatizing each word (i.e., converting each word into its root word). We then build a corpus of the documents using the text of the 147 patients. For the BoW Classic method, we then count the frequency of each unigram and bigram that appears in each patient text. That is, if a word or a pair of words appear in the notes for a patient, then that phrase appears in the patient vector. To further reduce dimensionality, we extract the final features by performing singular value decomposition (SVD). BoW TF-IDF is an advanced variant of BoW Classic that in addition to the steps outlined for BoW Classic controls for words that appear very frequently in texts but have low predictive power.

BERT, DistilBERT, ClinBERT, and Longformer, on the other hand, are cutting edge, deep-learning powered methods that have shown improved performance in numerous applications^19,21,43^. BERT^44^, the forerunner of these methods, leverages transfer learning from attention-based transformers, enabling it to understand the complex and evolving meaning of a word in a text in relation to all surrounding words, rather than strictly focusing on the interpretation of a word from reading text left to right. This model was pre-trained on the vast English Wikipedia and the BooksCorpus. DistilBERT^45^, a variant of BERT, is considered to be faster to pre-train, while still being able to retain most of the understanding capabilities. Similarly, ClinBERT^19^ uses the same conceptual framework as BERT but is instead tuned specifically to the medical context by training on clinical notes in the MIMIC database. Alternatively, Longformer^46^ is modified from BERT such that it can process longer text sequences. For these methods, we split the raw data into chunks of equal size that each method can process (chunks of size ∼512 tokens for BERT, DistilBERT, and ClinBERT and of size ∼4096 tokens for Longformer), apply the method onto each chunk, and concatenate the features. Similar to the BoW methods, we then additionally perform SVD to reduce the feature dimension.

We denote this set of features extracted by a NLP method as “AFG features”.

### 2.7 Outcome Prediction Machine Learning Models

To assess the value of the CFG process in extracting features to predict prostate cancer recurrence, we compare the performance of machine learning (ML) models trained and tested using different feature sets. In particular, we compare the performance of models with three different feature sets: 1) baseline features only (‘Baseline’ model); 2) baseline features and AFG features (‘Baseline + Automated NLP’ model); 3) baseline features and CFG features (‘Baseline + Patient-Level Clinician’ model). The Baseline model incorporates minimal clinician reasoning to create relevant features from structured data sources and acts as a control to compare the performance of the latter two models. Supplement B further details the features in each of the different models.

As the main outcome prediction ML model, we train LASSO models^47,48^ with these different feature sets, using 3-fold cross validation to tune the penalty parameter λ and the area under the receiver operating characteristic curve (AUC-ROC) as the performance metric. We choose a LASSO model as the main model because it is a linear model, allowing it to be more interpretable compared to other models. Furthermore, LASSO only has one parameter to tune (unlike other models such as XGBoost or deep learning), and given our small sample size, we want to ensure we do not overfit the model to our data. We repeat the training and validation process 50 times, each time using a different seed to split the data during cross validation (CV), and impute missing values for each model separately using SVD-imputation. We then compare the distribution of the AUC-ROC of the models across the 50 seeds.

We perform sensitivity analyses on the number of cross validation folds (considering 5-fold cross validation), the imputation method of missing values (considering median and multiple imputation by chained equations (MICE)), the performance metric (considering the area under the precision recall curve), and the machine learning model (considering k-Nearest Neighbors tuned for the number of neighbors, and a random feature model^49^ with ReLU activation centered at 0 tuned for a Ridge penalty parameter^50^, which mimics a two layer neural network with 1000 features in the hidden layer where first layer weights are generated from a multivariate standard normal distribution).

We also explore which features are most frequently used by the ‘Baseline’ model and ‘Baseline + Patient-Level Clinician’ model to analyze feature importance (detailed description of the process is in Supplement C).

## 3. Results

### 3.1 Comparison of BF Prediction Value

Figure 3 shows the distribution of the AUC-ROC for the ‘Baseline’, ‘Baseline + Automated NLP’, and ‘Baseline + Patient-Level Clinician’ model for the 50 iterations of the main outcome prediction ML model. For the ‘Baseline + Automated NLP’ model, we show the performance using features generated from progress notes with the NLP method Longformer as it outperforms the other five NLP methods for the main outcome prediction ML model (see Supplement D for further details).

**Figure 3:**
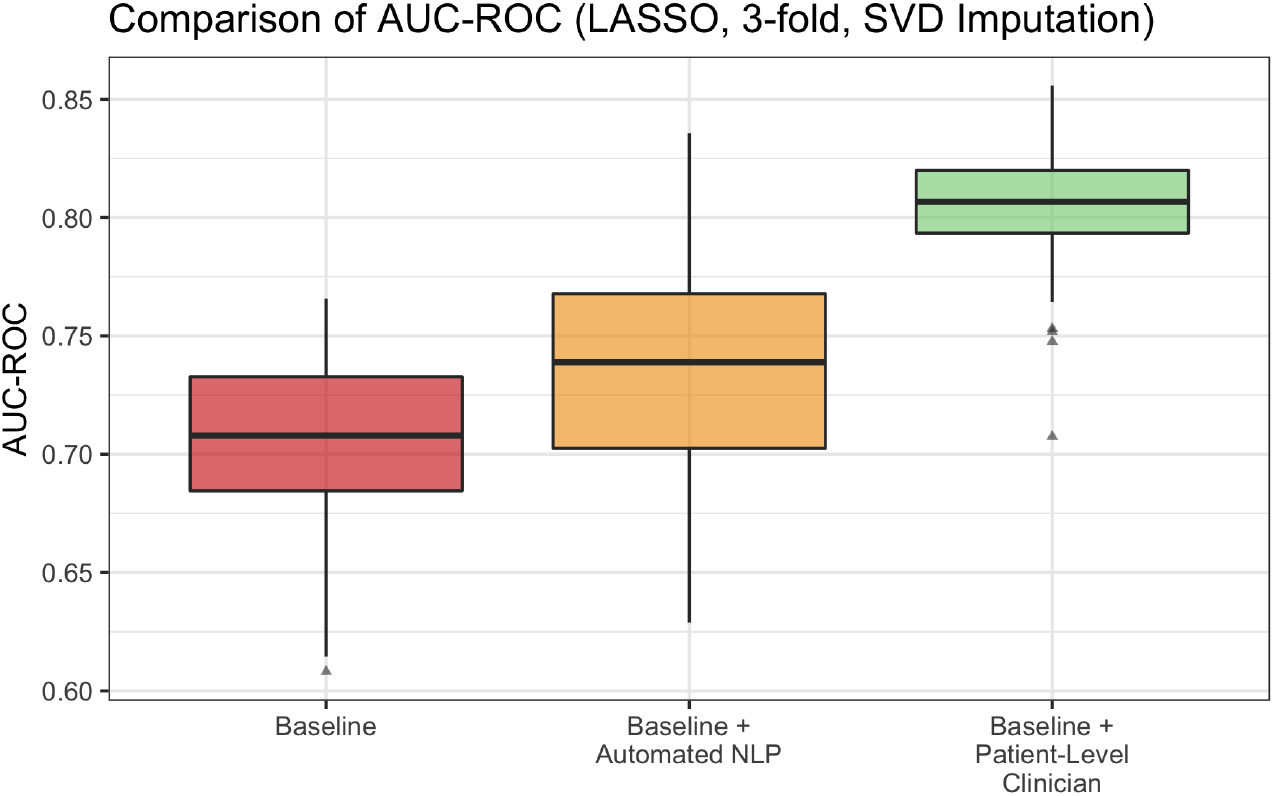
Comparison of the distribution of the AUC-ROC for the ‘Baseline’ (minimal clinical expertise), ‘Baseline + Automated NLP’ using the NLP method Longformer, and ‘Baseline + Patient-Level Clinician’ model for 50 iterations of the main model (LASSO, 3-fold cross validation, SVD imputation for missing values).

Comparing the distribution of the ‘Baseline + Patient-Level Clinician’ model (mean: 0.80) to the distribution of the ‘Baseline’ model (mean: 0.70), the former distribution dominates the latter, evidencing that there are features predictive of prostate cancer recurrence in patient notes. This is in line with Hsu et al.’s work^52^ on the readmission prediction task and other studies^17,51^. Further comparing the distribution of the ‘Baseline + Patient-Level Clinician’ model to the ‘Baseline + Automated NLP’ model (mean: 0.74), we conclude that the patient-level CFG process of leveraging extensive clinical expertise to identify and create patient-level features is able to extract more signal from the progress notes, compared to the AFG method via NLP. This highlights the value of utilizing the CFG process to generate features to predict prostate cancer recurrence more accurately.

To ensure robustness of our results, we perform extensive sensitivity analyses on the data pre-processing and machine learning model characteristics. We find that the conclusions are consistent across sensitivity analyses on the number of folds used to cross validate and tune the parameter of the LASSO model (Figure 4a), the imputation method applied to impute missing values (Figure 4b), the ML model trained (Figure 4c), and the performance metric used to train and evaluate the models (Figure 4d). Additionally, the results are qualitatively unchanged for the sensitivity analysis on the alternative BF definition, the timing of the biopsy report that contains the 12 systematic cores, and the removal of the features race, ethnicity, and language, as seen in Supplement E. As in the main outcome prediction ML model, the results of the sensitivity analyses firstly show that the inclusion of CFG features generated from progress notes improves the model performance in predicting prostate cancer recurrence, compared to only including the baseline features. And secondly, the CFG process is able to realize a higher level of performance compared to the AFG process, emphasizing the value of in-depth specialist reasoning in generating features.

**Figure 4:**
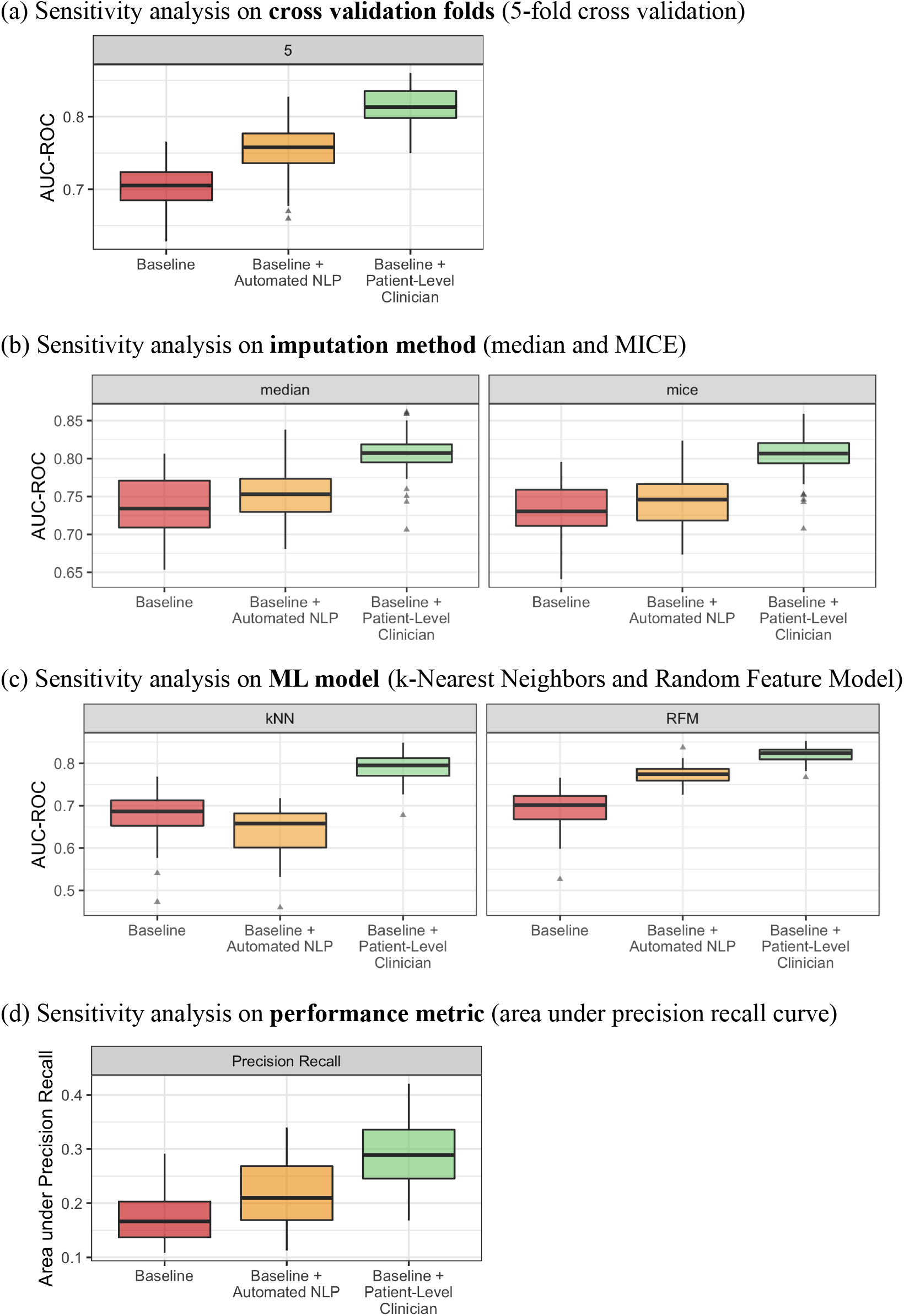
Sensitivity analysis results for the ‘Baseline’, ‘Baseline + Automated NLP’, and ‘Baseline + Patient-Level Clinician’ model, varying the number of cross validation folds (Figure 4a), imputation method (Figure 4b), ML model (Figure 4c), and performance metric (Figure 4d).

### 3.2 Feature Importance Analysis

For the ‘Baseline’ model, the most frequently non-zero features are the maximum pre-treatment PSA level and the Charlson Comorbidity Index, with the fraction of time that they are non-zero across the seeds being 0.72 for both. Given the ‘Baseline’ model does not have a lot of detailed information pertaining to the cancer, as those traits are in biopsy reports and clinical notes, its top non-zero features illustrate that most of its predictive power is retrieved from features that influence a patient’s general health. For the ‘Baseline + Patient Level Clinician’ model, the fraction of time that any systematic core grade group, percent Gleason pattern 4/5, and percent involved coefficient is non-zero is 0.80, 0.56, and 0.42, respectively, comprising the majority of non-zero coefficients in the model across the seeds. This analysis continues to highlight that the ‘Baseline + Patient-Level Clinician’ model gains predictive power from the specialist curated features found in progress notes that require the patient-level CFG method to identify and manual patient-level review to extract.

Supplement C shows the fraction of time that a feature is non-zero for all features included in the ‘Baseline’ and ‘Baseline + Patient-Level Clinician’ model for reference.

## 4. Discussion

Our analyses illustrate that the patient-level CFG approach developed, while time-intensive, greatly enhances the ML outcome prediction model and achieves a high performance in predicting the actionable outcome of 5-year prostate cancer recurrence. This method uses in-depth clinical expertise to identify clinically relevant concepts and to translate them into features that can be retrieved from structured data and patient-level progress notes. In contrast, in our study, an automated approach for feature generation that uses minimal clinical expertise and NLP methods to extract features is unable to retrieve as much signal. The AFG method lacks the patient-level human reasoning and specialist expertise required to extract features that emulate clinical knowledge to accurately predict prostate cancer recurrence. We find that these conclusions are robust to various sensitivity analyses including the choice of ML model, the tuning and imputation of missing values methods, and the performance metric. We further evaluate the key predictors in the ‘Baseline + Patient-Level Clinician’ model and find that features identified through the CFG process and manually curated from progress notes frequently exhibit non-zero coefficients in the ML outcome prediction model, signaling their value in and requirement to accurately predict prostate cancer recurrence.

The results of our study emphasize the need to research new objectives that assess a collective of in-depth clinical expertise and automated algorithms together for feature generation. Automated feature generation is unable to extract the entire signal from structured data and progress notes to predict prostate cancer recurrence and may suffer from biases. Additionally, these methods may not be generalizable from institution to institution. While there are many reasons for this, one contributor is that these methods use proxies rather than main underlying patient conditions that contribute to the outcome to create features. However, automated methods are easily scaled making them more accessible to gain traction in clinical practice. In contrast, patient-level clinician feature generation leads to improved predictive performance and likely does not exhibit the shortcomings of generalizability. In return, however, this method has the looming challenge of scaling at large as it requires the time-intensive manual retrieval of features from progress notes.

Our study therefore provides a proof of concept that there is a need to build a method that leverages the advantages of the two different approaches to retrieve CFG features in a scalable fashion, actively informed by the knowledge of clinicians. This retrieval could build on previous work exploring feature extraction methods^53–57^. Another first step to this could be to streamline the retrieval of the CFG features identified in progress notes into routine practice (e.g., requiring pathologists to fill out the grade group and percent involved per systematic core in a tabular fashion) to make them structured features in EHR data.

More broadly, our findings echo a well-known concept in ML and statistics: in a small sample regime, ML models typically suffer from overfitting and too little training data, decreasing their performance, and a Bayesian approach in which prior information is provided to the model can help boost its performance. Our results align with this notion as they illustrate the benefit of leveraging in-depth clinical expertise in this small sample regime to guide the feature generation process of a ML model. We recognize that the smaller sample size may hinder the performance of AFG when generating features from the progress notes via NLP methods that require a large corpus to tune accurately. However, to our knowledge, there does not exist a large de-identified corpus of progress notes for prostate cancer patients to better tune the NLP methods presented in our study, and we explored numerous different types of NLP methods in this work and find their performance to be inferior to the patient-level CFG approach. Additionally, given the requirement to manually retrieve the majority of the CFG features, it is difficult to easily extend this study to a larger sample size. This obstacle motivates the valuable future work discussed above to study how we can automate the extraction of the CFG features across multiple healthcare institutions.

One limitation of our work is that all patients studied are treated at the same healthcare institution. While multi-institution studies are important, our work hopes to address institutional discrepancies by incorporating clinical expertise in model building, which can make them more generalizable, rather than solely depending on features that can be programmatically extracted from available raw data, which may bias the model to rely on institution-specific artifacts.

Additionally, studies that analyze progress notes suffer from the limitation that notes can exhibit variability in writing style and description. Given the CFG features are extracted from notes written by various oncologists over the span of at least 10 years, there could be discrepancies in how a patient’s disease state is classified and described throughout. For example, there could be differences in the clinical staging standard that is used. Also, as highlighted in Section 2.5, the computation of the percent of the core involved at times requires human discrepancy in identifying the length of the core in the text and dividing accordingly, taking into consideration the different units used to record the information. To ensure high-quality data, for notes where identification of a feature was unclear, our team consulted experts, such as pathologists, who write these notes first-hand and they advised us on the best way to interpret the text and ultimately compute the feature of interest.

As the availability of detailed, patient-level information in EHR data grows, it is becoming crucial to create methods that can emulate human-reasoning to extract relevant predictors. Our study underscores the importance of leveraging progress notes for predicting prostate cancer recurrence and researching efficient integration of in-depth clinical expertise into feature generation for clinical risk prediction (e.g., designing a platform involving humans and AI).

## Data Availability

Summary statistics of the data are available upon reasonable request to the corresponding author. The raw patient data includes PHI and is thereby protected by privacy laws and cannot be shared.

## Supplement

### A. CFG Concept Abstraction

| **Clinical Concept** | **Features Representative of Concept** |
| --- | --- |
| Tumor Volume | Percent involved and percent Gleason pattern 4/5 per systematic core; maximum percent Gleason pattern 4/5, mean percent Gleason pattern 4/5, mean percent involved, and percent of positive regions; clinical T stage |
| Tumor Grade | Percent Gleason pattern 4/5 and grade group per systematic core; maximum percent Gleason pattern 4/5 and mean percent Gleason pattern 4/5; maximum pre-treatment Gleason score and corresponding grade group |
| Extraprostatic Extension | Clinical T stage |
| Regional Metastasis | Clinical N stage |

### B. Feature Sets and Main Outcome Prediction ML Model

To quantify the value of the CFG process in extracting features to predict prostate cancer recurrence, we compare the performance of machine learning (ML) models trained and tested on different feature sets. Table B.1 below outlines the three distinct feature sets, along with a detailed list of the features included in each. To extract the percent Gleason pattern 4/5, percent involved, and grade group per systematic core, we read the note to determine whether the feature is explicitly stated (it is typically found in the ‘diagnosis’ section if it is explicitly stated) or, if not, read the entire note in greater depth and, if necessary, compute the feature from the information present (e.g., compute the percent involved using the different length measurements provided). If the percent Gleason pattern 4/5 per systematic core is not explicitly stated, we use a clinical proxy (100% of the fraction of the core involved if both the Gleason primary and Gleason secondary score is 4 or 5; 75% of the fraction of the core involved if only the Gleason primary pattern is 4 or 5; and 25% of the fraction of the core involved if only the Gleason secondary pattern is 4 or 5). The main outcome prediction ML model considered is a LASSO model with 3-fold CV and the AUC-ROC as the performance metric. To impute missing values, we use SVD-imputation iterated 500 times with a rank of three.

**Table B.1:** List of the features in each feature set used to train and test various ML models.

| Model | Features | List of Features | No. of Features |
| --- | --- | --- | --- |
| **Baseline** | Baseline features | Race (White vs. non-White), ethnicity (Hispanic vs. non-Hispanic), age at treatment, marital status (with vs. without partner), primary language (English vs. other), maximum pre-treatment PSA level, Charlson Comorbidity Index three years prior to start of treatment | 7 |
| **Baseline + Automated NLP** | Baseline features + AFG features | See above list of baseline features + NLP Longformer generated features | 27 |
| **Baseline + Patient-Level Clinician** | Baseline features + CFG features | See above list of structured features + Clinical T and N stage; percent Gleason pattern 4/5, percent involved, and grade group per systematic core; maximum pre-treatment Gleason score (Gleason primary and Gleason secondary score) and corresponding grade group; mean percent Gleason pattern 4/5, maximum percent Gleason pattern 4/5, mean percent involved, and percent of positive regions, computed from all cores sampled in the biopsy, grouping cores by similar regions first | 52 |

### C. Feature Importance Analysis

Given the enhanced performance of the ‘Baseline + Patient-Level Clinician’ model, we further explore which features are most frequently used by the ‘Baseline’ and ‘Baseline + Patient-Level Clinician’ model in predicting cancer recurrence. To analyze this, for each seed and either the ‘Baseline’ or ‘Baseline + Patient-Level Clinician’ model features, we run the main outcome prediction ML model to determine the penalty parameter that leads to the highest AUC-ROC. We then analyze which features’ coefficient is non-zero in a LASSO model trained on two-thirds of patients, using this best penalty parameter. We then repeat this for 50 seeds and compute the frequency that each feature is non-zero across the repetitions.

For the ‘Baseline + Patient-Level Clinician’ model, the features of grade group, percent Gleason pattern 4/5, and percent involved per systematic core of the prostate each consist of 12 features corresponding to the 12 systematic cores. To analyze the feature importance, we group the 12 features for each of these into one category and consider the feature category to be leveraged by the model if any of the 12 systematic cores had a non-zero coefficient for that seed.

**Table C.1:**
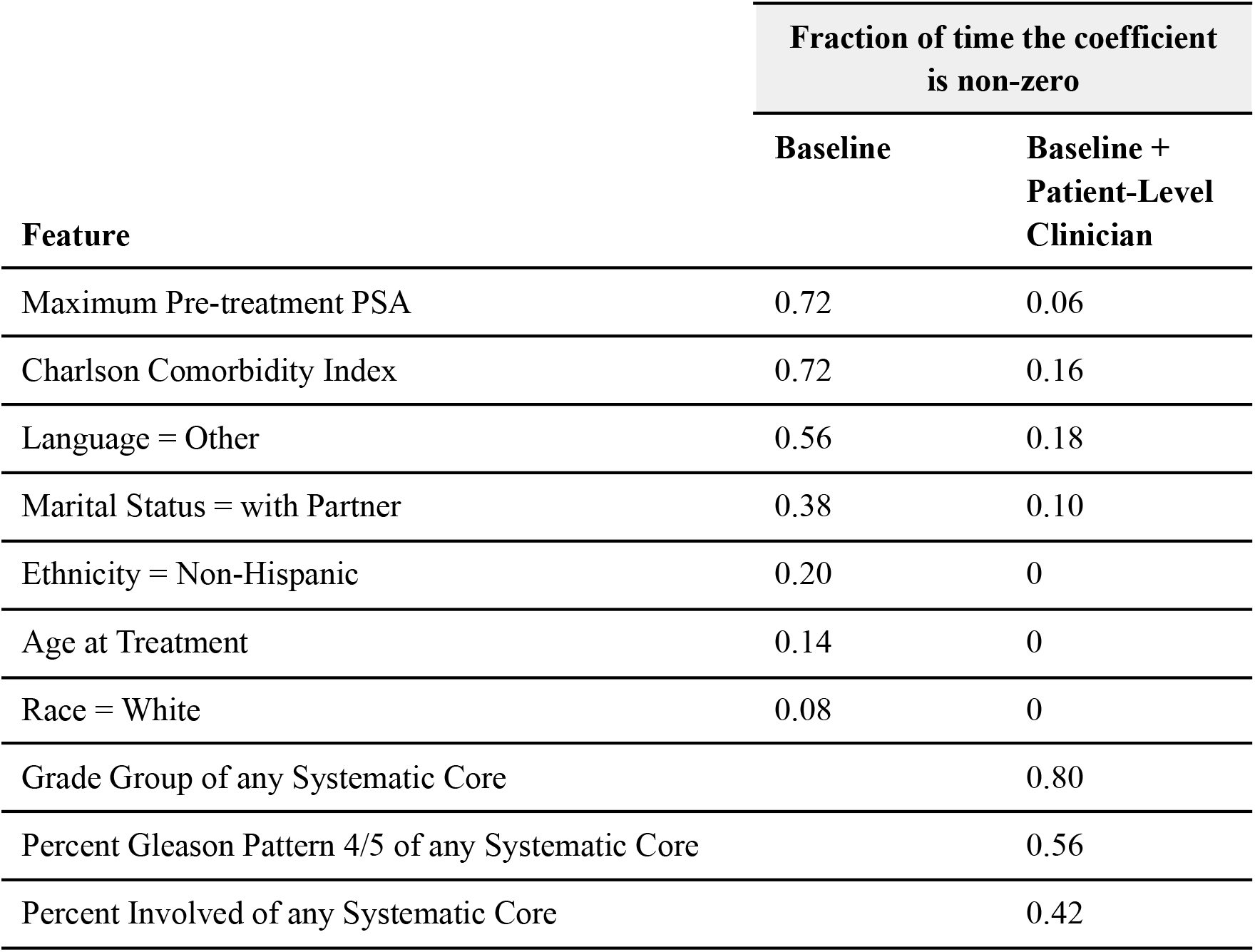
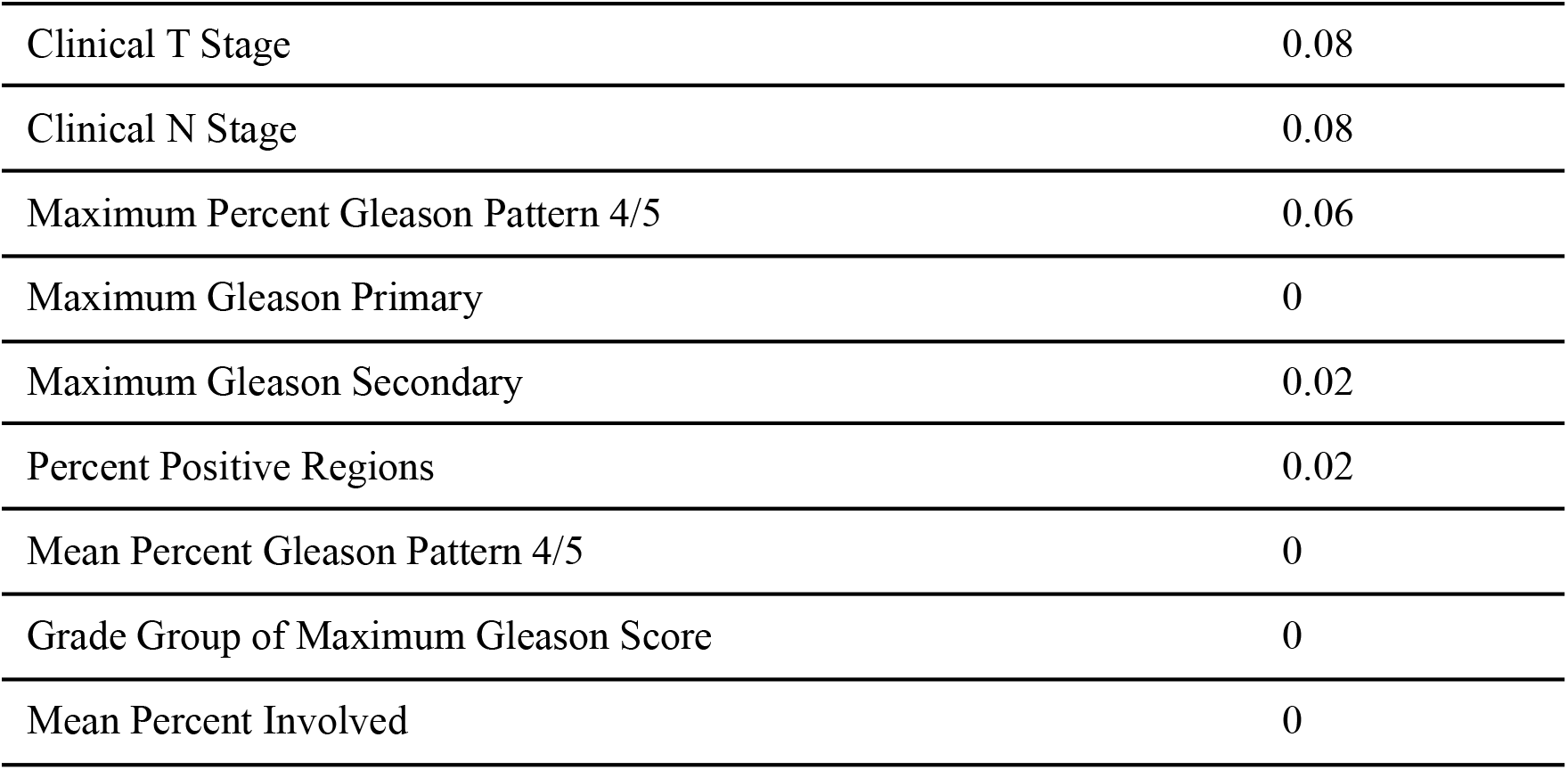
Fraction of time the coefficient of the feature is non-zero for the ‘Baseline’ and ‘Baseline + Patient-Level Clinician’ model, across 50 seeds of the main outcome prediction ML model.

### D. NLP Techniques

In our main results, we compare the performance of the ‘Baseline’ and ‘Baseline + Patient-Level Clinician’ model to the performance of the ‘Baseline + Automated NLP’ model which generates features from progress notes using the NLP method Longformer. We choose this method as it outperforms the performance of the other five NLP methods we consider: BoW Classic, BoW TF-IDF, DistilBERT, BERT, and ClinBERT. To get the performance for each of these methods, we apply each method separately on the concatenated progress note texts as described in Section 2.6. We then visually examine the singular values in log scale for each and select the top left singular vectors, based on the gap found, as the patient features in combination with the baseline features. Figure D.1 illustrates that the NLP method Longformer outperforms the other five NLP methods when applying the main outcome prediction ML model.

**Figure D.1:**
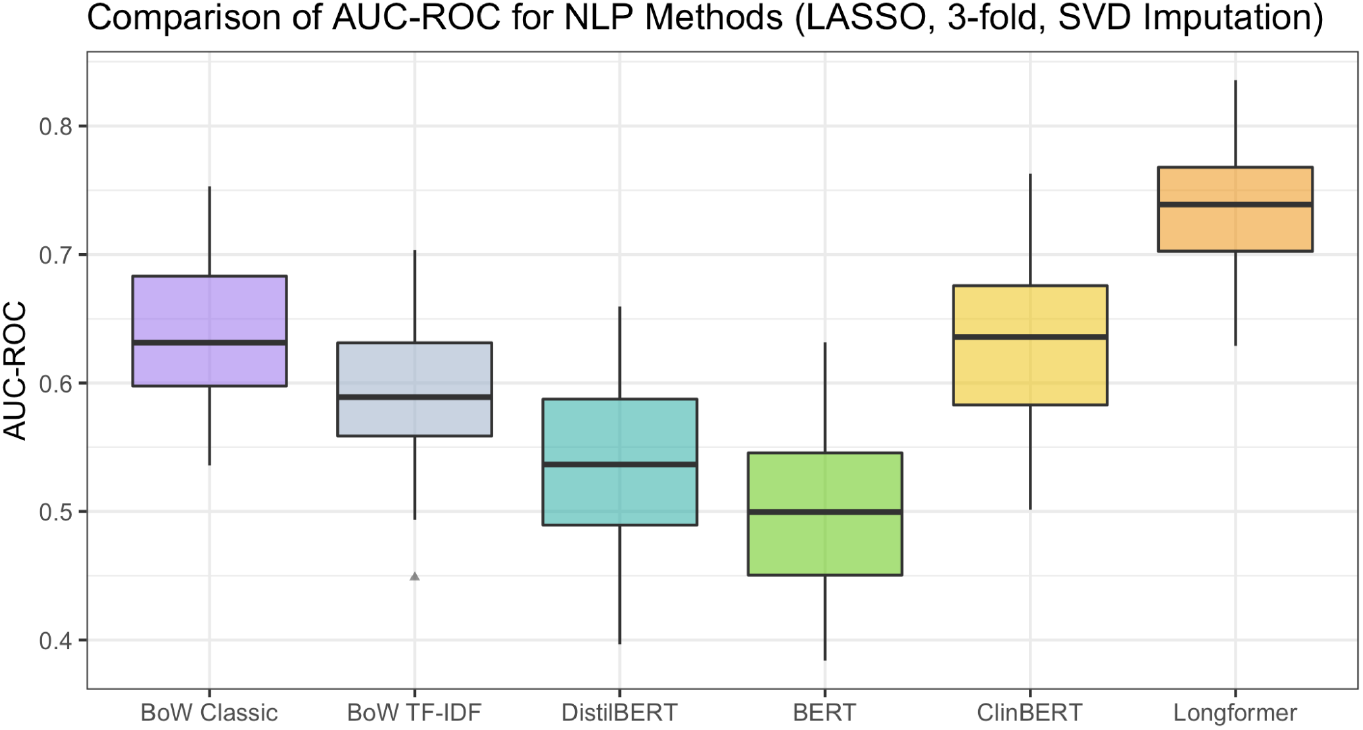
Comparison of the AUC-ROC for the six different NLP methods using the main outcome prediction ML model.

### E. Further Sensitivity Analyses

We complete sensitivity analyses varying the BF definition for prostatectomy patients, reducing the patient cohort determined by the timing of the biopsy report used to retrieve features for the 12 systematic cores, and removing the features of race, ethnicity and language.

Figure E.1a shows the results of the three main models using the main outcome prediction ML model (LASSO, SVD imputation, AUC-ROC for performance metric, 3-fold cross validation) when applying the BF definition for prostatectomy patients that uses the 0.2 ng/mL threshold described in Section 2.3. Figure E.1b similarly shows the results of the three main models when considering only patients who have a biopsy report with the 12 systematic cores recorded as their most recent biopsy report pre-treatment, reducing the sample size to 142 patients. In our paper, we allow the pre-treatment biopsy report from which we retrieve the features corresponding to the 12 systematic cores to be any pre-treatment biopsy report, and choose this approach to maximize our sample size. And, lastly, Figure E.1c shows the results of the three main models when removing the features race, ethnicity, and language, to ensure that our results are not reliant on these features and that the models are not using these features in any biased fashion. For both the change in BF definition, patient cohort selection, and feature selection, we see that results are qualitatively unchanged.

**Figure E.1:**
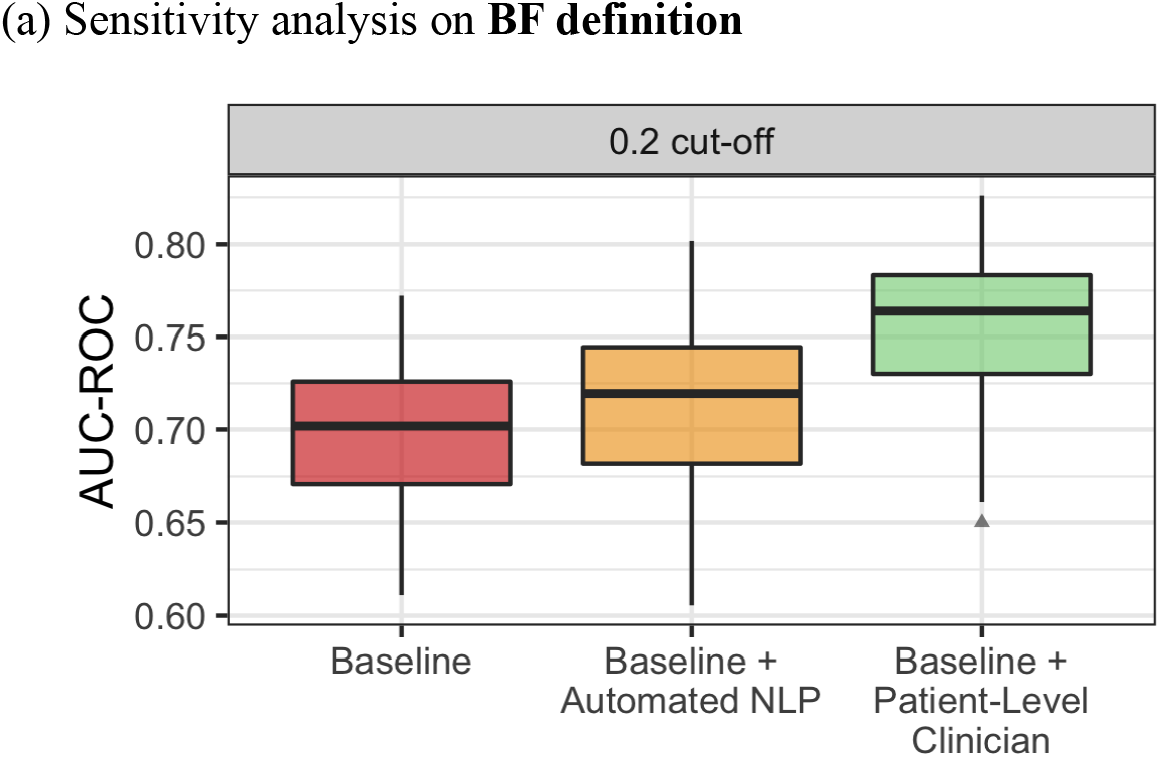
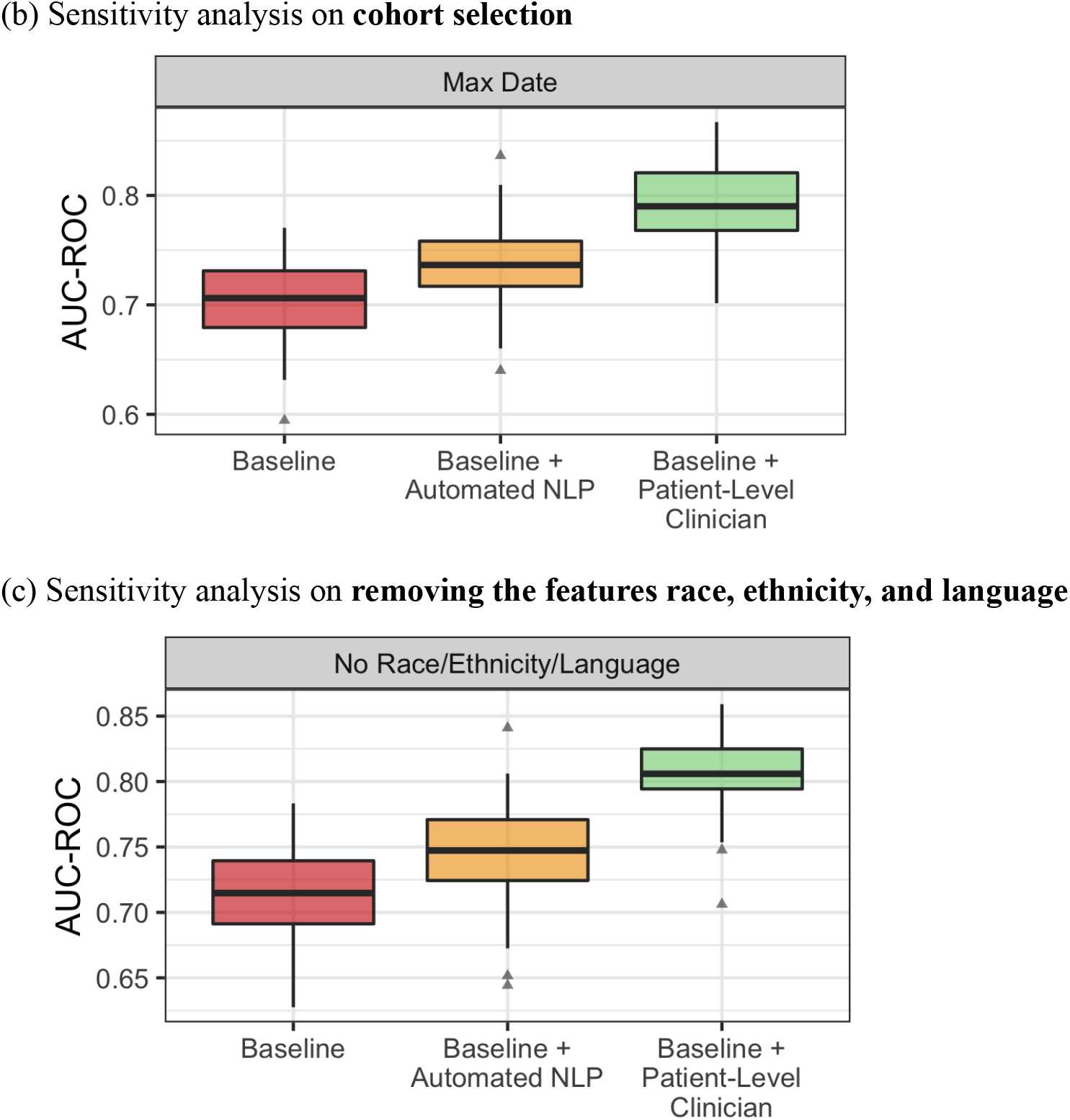
Sensitivity analysis results for the three models, varying the BF definition for prostatectomy patients (Figure E.1a), the cohort selection (Figure E.1b), and the inclusion of the race, ethnicity, and language feature (Figure E.1c).

